# Internet-based health survey on loneliness and sleep-related problems among the working-age population in Japan during COVID-19

**DOI:** 10.1101/2021.11.07.21266001

**Authors:** Hirofumi Tesen, Yusuke Konno, Seiichiro Tateishi, Ayako Hino, Mayumi Tsuji, Akira Ogami, Masako Nagata, Keiji Muramatsu, Reiji Yoshimura, Yoshihisa Fujino, for the CORoNaWork Project

## Abstract

**Background:** The coronavirus disease 2019 (COVID-19) pandemic has been linked to a rise in loneliness. Loneliness is associated with sleep-related problems, which in turn can be a risk factor for various psychiatric disorders. However, it is unclear whether loneliness is linked to sleep-related problems during the pandemic. Here, we studied the association between loneliness and sleep-related problems during the COVID-19 pandemic in Japan.

**Methods:** A total of 33,302 individuals who indicated they were employed were surveyed online. The survey responses of 27,036 participants were analyzed. Odds ratios (ORs) were estimated using univariate and multiple logistic regression analyses.

**Results:** Of those analyzed, 2,750 (10.2%) experienced feelings of loneliness. Further, sleep-related problems were significantly more common among those who felt lonely both in the short term (more than 3 days) and the long term (more than 3 months). The OR was much weaker after adjusting for factors related to interpersonal connections, such as family and friendships, than after adjusting for factors related to socioeconomic status.

**Conclusions:** Loneliness may be a risk factor for sleep-related problems in the COVID-19 pandemic. Having connections with family and friends may have a moderating effect on the occurrence of sleep-related problems.

## Introduction

Since the first confirmed cases of coronavirus disease 2019 (COVID-19), the disease has become a major infection risk around the world. Additionally, the associated pandemic has posed numerous other public health challenges such as loneliness.^2,13^ Physical distancing and curtailing outings and opportunities for socializing are some of the recommended measures for preventing infection. Specifically, governing bodies around the world have requested the public to refrain from going out as much as possible, conduct work and leisure activities at home, and refrain from socializing with those other than family members as much as possible. These recommendations are being linked to increased loneliness. One study reported that 35% of residents who experienced lockdown in China had psychological distress, while another demonstrated that 45% of adults in the US had anxiety and stress.^24,25^ The circumstances of those who experience loneliness have been worsened by the pandemic.^10^ Further, individuals with heightened stress of anxiety and loneliness have poorer sleep quality.^33^

Even before COVID-19, loneliness was an emerging public health issue. Researchers had begun to explore the possibility that loneliness may be a trigger for public health intervention for all generations.^11^ According to previous studies, 10–40% of the population experienced loneliness and isolation.^19,32^ While isolation refers to a lack of social interaction, loneliness is linked to subjective feelings. Although different, they are related, with isolation and loneliness shown to adversely affect health through both common and different pathways.^23^ Loneliness is associated with lower subjective health and lower quality of life, and exacerbates signs of depression.^18^ It is also a risk factor for suicide and dementia.^1,8,16^

In particularly, loneliness is associated with sleep-related problems, which in turn can be a risk factor, precursor, or accompanying symptom of various psychiatric disorders. In the COVID-19 pandemic, loneliness has been identified as a major risk factor for insomnia.^15^ A study in Japan on patients who visited a psychiatric clinic during the pandemic demonstrated a link between loneliness and earlier bedtime and increased sleep duration.^27^ Other reports suggest that sleep disorders are on the rise during the pandemic. ^29^

Despite reports of an increase in people experiencing loneliness and isolation during the COVID-19 pandemic, the relationship between loneliness and sleep-related problems is unclear. Here, we studied the relationship between loneliness and sleep-related problems during the COVID-19 pandemic in Japan.

## Methods

### Study design and participants

The present analysis forms part of the Collaborative Online Research on the Novel-Coronavirus and Work (CoroNaWork) Project, a cross-sectional study conducted between December 22 and 26, 2020, that used Internet-based surveys to probe the health of Japanese employees during the COVID-19 pandemic. A full description of the protocol is provided elsewhere.^9^ The survey was performed on individuals with an employment contract. Individuals whose response time was extremely short, height was below 140 cm, weight was below 30 kg, or provided conflicting answers to the same question were excluded. Out of 33,302 participants, responses from 27,036 were analyzed.

This study was conducted with the approval of the Ethics Committee of the University of Occupational and Environmental Health (approval number R2-079). Informed consent was obtained through a form on the survey website.

### Assessment of loneliness

We used a questionnaire to assess participants’ loneliness. The questionnaire asked how often the participants had felt lonely during the last 30 days. Those who answered “never” or “a little” were grouped as feeling no loneliness. In contrast, those who answered “sometimes,” “usually,” or “always” were grouped as feeling loneliness.

### Assessment of sleep

We used a questionnaire to assess participants’ sleep status. The questionnaire asked three questions. The first asked whether participants were getting enough sleep. The second asked whether they had experienced any trouble sleeping for more than 3 days. The third asked whether they had experienced any trouble sleeping for more than 3 months. Participants answered yes or no to these questions.

### Other covariates

For analysis, we treated the following as confounding factors: age, sex, marital status, equivalent income, education, smoking, job type, number of employees in the workplace, cumulative incidence rate of COVID-19 in the prefecture of residence, lack of friends to talk to, lack of acquaintances to ask for favors, lack of people to communicate with through social networking sites, family time and solitary eating. Additionally, we used the cumulative incidence of COVID-19 in the prefecture of residence in the month prior to the survey as a community-level variable. These data were taken from the websites of public institutions.

### Statistical analysis

Odds ratios (ORs) were estimated using univariate and multiple logistic regression analyses. Loneliness was treated as an independent variable and the presence of sleep-related problems as a dependent variable. To determine the association between loneliness and sleep problems, we constructed two multivariate models. In model 1, we adjusted for age, sex, marital status, equivalent income, education, smoking, alcohol consumption, job type, number of employees in the workplace and cumulative incidence rate of COVID-19 in the prefecture of residence. In model 2, we additionally adjusted for lack of friends to talk to, lack of acquaintances to ask for favors, lack of people to communicate with through social networking sites, family time and solitary eating.

All analyses were conducted using Stata (Stata Statistical Software: Release 16. College Station, TX: StataCorp LLC.), with p<0.05 indicating statistical significance.

## Results

Table 1 summarizes the general characteristics of the 27,036 participants included in the study. Of those analyzed, 2,750 (10.2%) experienced feelings of loneliness. Age, region, occupation, and income were comparable between those who felt lonely and those who did not. On the other hand, those who reported feeling lonely were more likely to be unmarried, divorced or bereaved.

Table 2 summarizes the ORs of loneliness associated with sleep-related problems as estimated by the logistic model. We found a significant association between loneliness and the presence of sleep-related problems evaluated using the question “Do you get enough sleep?” The age-sex adjusted OR was 2.64 (95% CI 2.43–2.87). The association remained significant after adjusting for confounders in model 1 (OR=2.58, 95% CI 2.37–2.80) and model 2 (OR=2.05, 95% CI 1.89–2.24). A significant association was also observed between loneliness and the presence of short-term sleep-related problems based on the question “Have you had any trouble sleeping for more than 3 days?” The age-sex adjusted OR that participants who felt lonely had sleep-related problems was 3.63 (95% CI 3.35–3.94). The association was likewise significant in model 1 (OR=3.53, 95% CI 3.25–3.83) and model 2 (OR=2.95, 95% CI 2.71–3.22). Further, we also observed a significant association between loneliness and the presence of long-term sleep-related problems based on the question “Have you had any trouble sleeping for more than 3 months?” Among those who reported feeling lonely, the age-sex adjusted OR for sleep-related problems was 3.59 (95% CI 3.31–3.90). Similarly, the association was significant in model 1 (OR=3.50, 95%CI 3.23–3.80) and model 2 (OR=2.87, 95%CI 2.64–3.13).

## Discussion

### Main finding of this study

We found that, during the COVID-19 pandemic, sleep-related problems were significantly more common among those who felt lonely both in the short term (more than 3 days) and the long term (more than 3 months). The OR of loneliness associated with sleep-related problems was much weaker when adjusted for factors related to interpersonal connections, such as family and friendships, than when adjusted for factors related to socioeconomic status. This suggests that having connections with family and friends has a moderating effect on the occurrence of sleep-related problems.

### What is already known on this topic

About 10% of participants in this study felt lonely. To our knowledge, this is the first large-scale study to investigate loneliness in a working-age population in Japan. According to a previous Japanese study, the percentage of individuals experiencing loneliness among those aged 65 and above who were living with a spouse only, living with children, and living alone was 17.7%, 18.5%, and 37.3%, respectively.^26^ The lower incidence of loneliness in the present study may reflect the fact that working-age individuals more actively participate in society through work, and are in the early stages of marriage and raising children. However, we found that workers who were unmarried, divorced, or had lost a partner; had no neighbors or friends to talk, ask for favors, or communicate with on social networking sites; had little time to spend with family, or ate meals alone tended to feel lonely despite working.

Our analyses showed that those who felt lonely were more likely to have sleep-related problems. These results are consistent with those of previous studies. One report found that pandemic-related loneliness, anxiety, and depression led to insomnia, which is more pronounced among women and inner-city residents.^29^ Another report also showed that pandemic-related loneliness and anxiety were associated with insomnia.^15^ Loneliness has been shown to be associated with sleep fragmentation and poor sleep quality.^17^ A study that adjusted for the effects of depressive symptoms suggested that the relationship between loneliness and insomnia cannot be explained by the comorbidity of depressive symptoms alone.^5^ When individuals experience loneliness and threats to the safety of the social environment, vigilance against social threats is enhanced and the brain remains alert during sleep.^3^ Those who maintain good relationships with others tend to choose healthy behavioral actions.^6^ Having social relationships and choosing healthy behaviors has been suggested to lead to good sleep quality.^4^

Loneliness and comorbid sleep problems may be interrelated in terms of their downstream consequences, which include psychiatric disorders, poor quality of daily life, and loss of work productivity. Sleep-related problems are a known risk factor for mental disorders.^30^ The effectiveness of cognitive behavioral therapy for comorbid insomnia in ameliorating psychiatric disorders has been reported.^12,20,22^ In addition, therapeutic interventions for insomnia have been shown to contribute to a reduction in social losses such as health problems, accidents, and lost productivity and mental illness.^21,31^ In addition to sleep, loneliness has also been shown to be associated with the risk of suicide attempts,^8^ dementia,^16^ alcohol use disorders and internal compliance problems.^14,19^ Caution is warranted when the two mix because sleep problems complicated by loneliness may also be associated with suicide risk, prognosis of mental disorders and difficulty introducing treatment.

### What this study adds

We found that having interpersonal connections with family and friends was effective in alleviating sleep-related problems in workers who felt lonely during the COVID-19 pandemic. The significant association of loneliness with sleep problems was true even after accounting for socioeconomic factors such as sex, age, and marriage. However, further adjusting for interpersonal connections with family and friends in model 2 led to a marked attenuation of the OR. To prevent spread of COVID-19 in Japan, the government has requested that people engage in physical distancing and refrain from going out. Self-isolation has been encouraged, for example, by performing work and leisure activities at home and refraining from interacting with those outside the family as much as possible. These requests may have brought the problem of loneliness to the surface for some workers. For those who live with their families, self-isolation allows them to spend more time and strengthen relationships with their kin. However, for workers who live alone or have no community ties outside of work, self-isolation may enhance the negative effects of loneliness.

Loneliness is a new challenge that workforces face as telecommuting and physical distancing have become recurring infection control measures. Although by nature, workers participate in society by working, some people continue to experience loneliness. As opportunities for direct communication with workers decrease through telecommuting and other means, it is necessary to identify workers experiencing loneliness to prevent downstream adverse effects. Workers who have no family or friends are particularly high-risk, especially during a widespread pandemic. In addition, sleep problems are common among workers. Since sleep problems complicated by loneliness may be associated with more severe mental disorders and difficulty introducing treatment, both loneliness and sleep should be evaluated in workers.

### Limitations of this study

First, because this study was conducted on Internet users, the degree to which the results are generalizable is unclear. To reduce bias, we sampled based on region, job type and prefecture according to the rate of infection. Second, whether or not participants felt lonely was determined using one question: “During the last 30 days, how often did you feel the following emotions?” Further studies using less subjective assessments of loneliness are needed to confirm our findings. Third, we were unable to assess the severity of sleep problems as we did not use the insomnia rating scale. Finally, because this was a cross-sectional study, we could not determine the temporal link between loneliness and sleep-related problems.

## Conclusion

Loneliness was found to be a risk factor for sleep-related problems during the COVID-19 pandemic. Our findings suggest that having connections with family and friends has a moderating effect on the occurrence of sleep-related problems. Workers who have no connections with family and friends are at high risk of sleep problems. Identifying workers who feel lonely and have reduced opportunities for direct communication during the pandemic may prevent adverse downstream effects. Given the pandemic is still ongoing, strategies are needed to manage loneliness and sleep-related problems.

## Data Availability

All data produced in the present work are contained in the manuscript

## Acknowledgements

This study was supported by a research fund from the University of Occupational and Environmental Health, Japan; General Incorporated Foundation (Anshin Zaidan); The Development of Educational Materials on Mental Health Measures for Managers at Small-sized Enterprises; Health, Labour and Welfare Sciences Research Grants; Comprehensive Research for Women’s Healthcare (H30-josei-ippan-002); Research for the Establishment of an Occupational Health System in Times of Disaster (H30-roudou-ippan-007), and scholarship donations from Chugai Pharmaceutical Co., Ltd.

Present members of the Collaborative Online Research on the Novel-coronavirus and Work (CORoNaWork) Project are: Dr. Yoshihisa Fujino (current chairperson), Dr. Akira Ogami, Dr. Arisa Harada, Dr. Ayako Hino, Dr. Chimed-Ochir Odgerel, Dr. Hajime Ando, Dr. Hisashi Eguchi, Dr. Kazunori Ikegami, Dr. Keiji Muramatsu, Dr. Koji Mori, Dr. Kyoko Kitagawa, Dr. Masako Nagata, Dr. Mayumi Tsuji, Dr. Rie Tanaka, Dr. Ryutaro Matsugaki, Dr. Seiishiro Tateishi, Dr. Shinya Matsuda, Dr. Tomohiro Ishimaru, Dr. Tomohisa Nagata, Dr. Yosuke Mafune, and Ms. Ning Liu, in alphabetical order. All of the members are affiliated with the University of Occupational and Environmental Health, Japan.

The authors acknowledge Dr. Yoshihisa Fujino, PhD, MD, and Yusuke Konno, MD, Department of Environmental Epidemiology, Institute of Industrial Ecological Sciences, University of Occupational and Environmental Health, Japan.

